# INTEGRATING INDIGENOUS PRACTICES WITH EVIDENCE-BASED CARE: A MIXED-METHODS INVESTIGATION OF BURULI ULCER TREATMENT DYNAMICS IN IMO STATE, NIGERIA”

**DOI:** 10.1101/2025.08.23.25334057

**Authors:** Chigozie Divine Onwuka, Oparaocha Evangeline Tochi

## Abstract

Buruli Ulcer (BU), caused by Mycobacterium ulcerans, is still a major public health concern in West Africa, particularly in rural populations such as Imo State, Nigeria. Despite the availability of effective WHO-recommended antibiotic regimens, a significant minority of affected people continue to use traditional and unconventional therapeutic methods, often delaying access to biomedical care. This interaction between traditional beliefs and contemporary medicine poses both obstacles and opportunity for improving BU management in endemic areas. This study analyzes the treatment dynamics of BU in Imo State by looking at how indigenous approaches are integrated with evidence-based medical treatments. It seeks to comprehend patient health-seeking behaviors, cultural views of the condition, and the impact of these variables on therapeutic outcomes. A concurrent mixed-methods strategy was used, with 400 laboratory-confirmed BU patients spread throughout three endemic local government districts in Imo State. Structured questionnaires were used to obtain quantitative data on treatment options, duration of delays, lesion categorization, and treatment results. To capture cultural and belief-driven influences on treatment decisions, qualitative data were collected through focus group discussions and in-depth interviews with patients, traditional healers, and healthcare personnel. 78% of the individuals sought indigenous or spiritual therapies first, with a median delay of 12 weeks before obtaining orthodox care. In this group, 82% developed to advanced lesions (Category II or III). Patients who received evidence-based care promptly had significantly greater healing rates (95%) and decreased complication rates (p<0.01). Strong cultural views (65%), skepticism in biomedical systems (16%), and economic limitations (30%) were the primary factors motivating unorthodox treatment preferences. Notably, several traditional healers expressed a readiness to work with mainstream health care institutions if properly engaged and taught. The incorporation of culturally sensitive health education and the involvement of trustworthy indigenous practitioners offer a viable strategy for promoting early detection and adherence to successful BU treatment in Imo State. Bridging traditional and biological techniques has the potential to considerably reduce disease load and enhance health outcomes in endemic situations. Policy innovations that focus on community participation and cross-sector collaboration are critically needed.

## 1. INTRODUCTION

Buruli Ulcer (BU) is a debilitating and necrotizing cutaneous infection caused by Mycobacterium ulcerans, primarily affecting individuals in remote, tropical, and impoverished settings. BU is recognized by the World Health Organization (WHO) as a neglected tropical disease (NTD) due to its low global visibility and the disproportionate burden it places on underserved populations (WHO, 2023). The infection typically begins as a painless nodule or papule and can progress to large ulcerative lesions if untreated, often resulting in permanent disability or disfigurement (Yeboah-Manu et al., 2021). In Nigeria, BU is an emerging public health concern with cases increasingly reported in several southern states, including Imo State (Onwuka, 2019). Despite the availability of effective antimicrobial therapies—most notably the WHO-recommended 8-week regimen of rifampicin and clarithromycin—the uptake of biomedical treatment remains suboptimal (Nigerian Ministry of Health, 2023). Patients often seek care late, primarily due to socio-cultural norms, financial barriers, and accessibility challenges (Garchitorena et al., 2019). Indigenous medical systems, which include herbal remedies, spiritual rituals, and consultation with traditional healers, are deeply embedded in the health-seeking behavior of many Nigerian communities. These practices are not merely alternative but are often the first line of care due to their accessibility, affordability, and cultural resonance. While these systems may provide psychosocial support, their lack of clinical efficacy in treating infectious diseases like BU poses a risk for delayed healing and increased morbidity (Adebayo et al., 2022). Given the complexity of treatment decision-making in resource-constrained settings, it becomes imperative to understand the interface between traditional and biomedical practices. This study aims to examine the treatment dynamics of BU in Imo State, with a particular focus on the drivers of indigenous care use, the clinical outcomes associated with delayed presentation, and the potential for integrating culturally competent strategies into conventional healthcare delivery. That can lead to long-term disability. Predominantly affecting children and impoverished adults in rural Africa, BU is caused by Mycobacterium ulcerans. Nigeria is among the endemic countries, with Imo State reporting recurrent cases. Although the WHO advocates an 8-week antibiotic regimen as first-line therapy, many patients rely on traditional healing practices, leading to treatment delays and complications. This study investigates the socio-cultural, economic, and clinical factors influencing treatment choices and explores possibilities for integrating indigenous and orthodox treatment models.

## 2. MATERIAL AND METHODS

### 2.1 Study Design

A concurrent mixed-methods design was adopted to capture both the quantitative prevalence and statistical associations related to BU treatment dynamics, as well as qualitative insights into patients’ beliefs and healthcare behaviors. This approach allowed for triangulation and deeper interpretation of results from different methodological lenses.

### 2.2 Study Area

Imo State, in the southeast region of Nigeria, was chosen as the study area. The estimated population of Imo State is 5,459,300 (NPC, 2022), making up 3.8% of Nigeria’s total population. Covering 5,100 square kilometers, the state is located between 4° 45’ and 7° 15’ North latitude and 6° 50’ and 7° 25’ East longitude as well. Geographically, Imo State is bordered to the north by Anambra State, to the east by Abia State, and to the south and west by Rivers State. Owerri, Okigwe, and Orlu are the three senatorial zones that make up the state, which also has 27 Local Government Areas (LGAs). The state’s three largest urbanized cities are Owerri, Okigwe, and Orlu, although the remainder of the population is spread among numerous rural areas.

The state’s economy is varied, with the main industries being small-scale manufacturing, tourism, sand mining, stone quarrying, fishing, trading, crop and animal farming, and government service. Numerous public and private schools, as well as a number of universities and colleges, make up the state’s well-established educational system. Imo State boasts 1,338 medical facilities dispersed among 27 LGAs. The Federal Medical Center, a public tertiary hospital in the state, acts as a referral hub for all other lower-level medical facilities.

The public hospital provides additional medical services through a number of private hospitals, such as the German Leprosy and Tuberculosis Relief Association (GLRA). Out of the 27 LGAs in Imo State, 3 Local Government Areas were chosen at random for this study (fig 2.1). These settlements are situated in regions where Buruli ulcer disease is endemic and at high risk. These three LGAs, Oguta, Obowo and Ikeduru were purposively selected due to their endemicity for Buruli Ulcer and their demographic representation of urban, peri-urban, and rural settings.

**Fig. 2_1.**
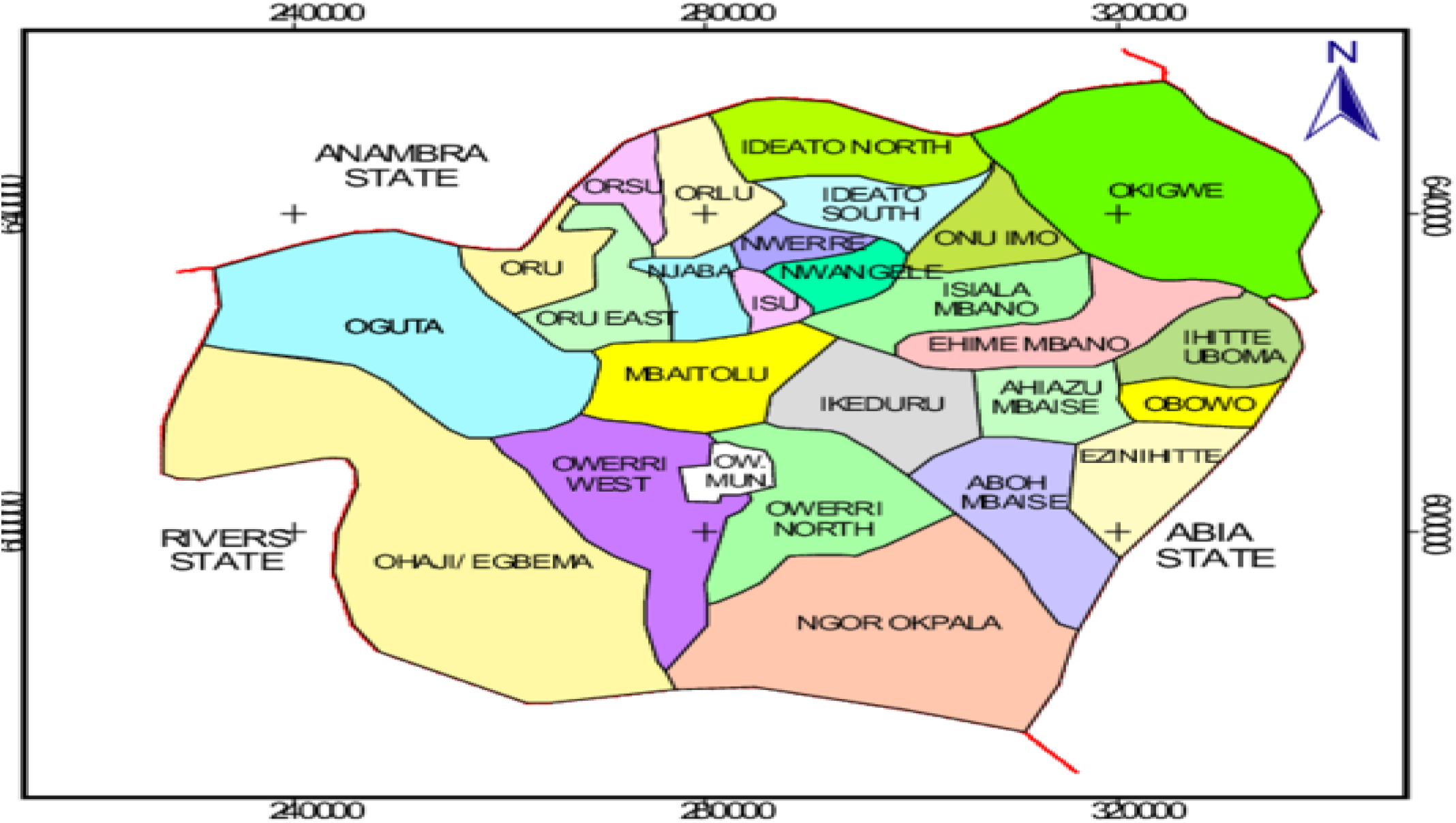
Sources *National population commission* (NPC, 2022)

### 2.3 Study Population

The study population included laboratory-confirmed BU patients, individuals, healthcare providers, and traditional healers who have direct or indirect experiences with BU treatment in selected communities of Imo State, Nigeria, particularly those within endemic Local Government Areas (LGAs) such as Oguta, Obowo and Ikeduru These communities were selected due to reported BU prevalence and the coexistence of orthodox medical facilities and indigenous/traditional health practices. The study population was intentionally designed to capture a wide range of perspectives, ensuring inclusivity of various stakeholders involved in BU diagnosis, treatment, and management.

### 2.4 Sampling and Sample Size Techniques

Given the mixed-methods nature of this study, a combination of probability and non-probability sampling techniques was employed to ensure both representativeness in the quantitative component and depth in the qualitative component.

#### Quantitative Sampling

Multi-Stage Sampling Approach: To select participants for the structured questionnaire survey, a multi-stage sampling technique was adopted:

##### Stage 1: Purposive Selection of LGAs

Three LGAs in Imo State, Oguta, Obowo and Ikeduru were purposively selected based on their known endemicity of Buruli ulcer, presence of indigenous healing systems, and accessibility.

##### Stage 2: Random Selection of Communities

Within each selected LGA, two communities were selected using simple random sampling (balloting). This ensured an even geographic spread and minimized selection bias.

##### Stage 3: Systematic Sampling of Households In each selected community

A systematic random sampling method was used to choose households. A sampling interval was determined based on the estimated number of households and the required sample size per community.

##### Stage 4: Selection of Respondents From each selected household

One eligible adult (≥18 years) was randomly selected for participation. Where more than one eligible respondent was present, balloting was used.

#### Qualitative Sampling

##### Purposive and Snowball Sampling

For the in-depth interviews and focus group discussions (FGDs), purposive sampling was employed to identify individuals with specialized knowledge or experience relevant to BU treatment dynamics, including:

a. Traditional healers/herbalists with history of treating skin lesions or BU
b. BU patients and survivors (both adherent and non-adherent to hospital treatment).
c. Healthcare workers (nurses, doctors, health officers) involved in BU care.
d. Local health administrators and program officers.

To identify hard-to-reach individuals, particularly traditional healers and BU survivors not currently linked to health facilities, snowball sampling was used where initial participants recommended others meeting the study criteria.

#### Sample Size Determination: Quantitative Component

The sample size for the quantitative survey was calculated using Cochran’s formula for sample size determination for populations greater than 10,000:

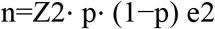

#### Where

Z = Z-value (1.96 for 95% confidence level)

p = estimated proportion of the population with awareness or experience of BU treatment (assumed at 0.5 for maximum variability)

e = desired level of precision (margin of error), set at 5% (0.05)

n =1.962⋅0.5⋅(1−0.5)0.052=3.8416⋅0.250.0025=0.96040.0025=384.16

Since the total population of adults in the selected communities was less than 10,000, the sample size was adjusted using the finite population correction formula:

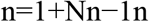

#### Where

N = estimated adult population in the selected communities (∼1000)

Applying the correction:

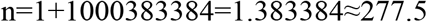

To account for non-response and incomplete questionnaires, a 10% contingency was added: Final sample size =278+ (10% of 278) =278+28≈306 respondents

### 3.5 Data Collection

The data collection phase of this mixed-methods study was carefully designed to obtain both quantitative and qualitative data relevant to the treatment dynamics of Buruli ulcer (BU) in Imo State, Nigeria. A convergent parallel design was employed, wherein both types of data were collected concurrently but analyzed separately and later integrated for triangulation and interpretation.

#### Quantitative Data Collection

Quantitative data were obtained through the administration of structured, interviewer-administered questionnaires. These questionnaires were designed to elicit information on: Socio-demographic characteristics (age, sex, education level, occupation, etc.), awareness and knowledge of Buruli ulcer symptoms and causes, treatment-seeking behaviors and healthcare utilization patterns, preferences for traditional versus orthodox treatment, barriers to accessing formal medical care, adherence to treatment protocols (among known BU patients).

#### Development and Validation of the Questionnaire

The questionnaire was developed based on previous BU-related studies (e.g., Ahorlu et al., 2013; WHO BU guidelines, 2020) and adapted to the local context through field pre-testing. It was reviewed by subject matter experts in public health, epidemiology, and medical anthropology to ensure content validity.

A pilot test was conducted in a non-selected but demographically similar community, and reliability was measured using Cronbach’s alpha, yielding a coefficient of 0.81, indicating high internal consistency.

### 2.6 Data Collection Procedure

Trained field enumerators fluent in both English and the local Igbo language conducted face-to-face interviews using printed or tablet-based forms. Data collection spanned ten weeks and covered both weekdays and weekends to increase response rates. Ethical considerations were strictly observed: informed consent was obtained, confidentiality was assured, and data were anonymized.

#### Qualitative Data Collection

To provide in-depth insights into cultural, social, and experiential dimensions of BU treatment, qualitative data were gathered using: Key Informant Interviews (KIIs), Focus Group Discussions (FGDs), Participant Observations (in selected traditional healing settings) Key Informant Interviews (KIIs): A total of 15 KIIs were conducted with: Community health workers, BU program officers, traditional healers/herbalists, BU survivors. These interviews explored themes such as: indigenous knowledge and interpretations of BU, treatment experiences, role of spiritual beliefs, stigma, and interactions with the healthcare system. Focus Group Discussions (FGDs): Eight FGDs (two per LGA) were held, each comprising 10–14 participants drawn from different community groups (youths, women, elderly, BU-affected households).

Discussions were moderated using a semi-structured guide, and themes explored included health-seeking behavior, trust in healthcare providers, and community perceptions of disease causation and treatment efficacy.

#### Data Recording and Management

All qualitative sessions were conducted in the local language or Pidgin English, recorded (with permission), and transcribed verbatim. Translations into English were done by bilingual experts and checked for accuracy. Field notes were also kept to capture non-verbal cues, context, and observational insights. .

### 3.7 Data Analysis

**Quantitative data** were analyzed using SPSS version 26. Descriptive statistics (means, frequencies, percentages) summarized socio-demographic variables and treatment patterns. Inferential analysis included Chi-square tests and logistic regression to identify predictors of late-stage presentation (Category III lesions).

**Qualitative data** were transcribed and coded using NVivo 12. Thematic analysis was employed to identify recurring patterns and concepts, organized under pre-defined and emergent themes related to cultural beliefs, economic barriers, and health system engagement.

## RESULTS

### 3.1 Quantitative

Out of the 306 respondents, 78% (n=256) reported initially consulting traditional healers before seeking orthodox medical treatment. The median delay before seeking biomedical care was 12 weeks (IQR: 8–16). Analysis of lesion severity at presentation revealed that 82% of patients who delayed treatment beyond 8 weeks presented with Category II or III lesions.

**Table 1.**
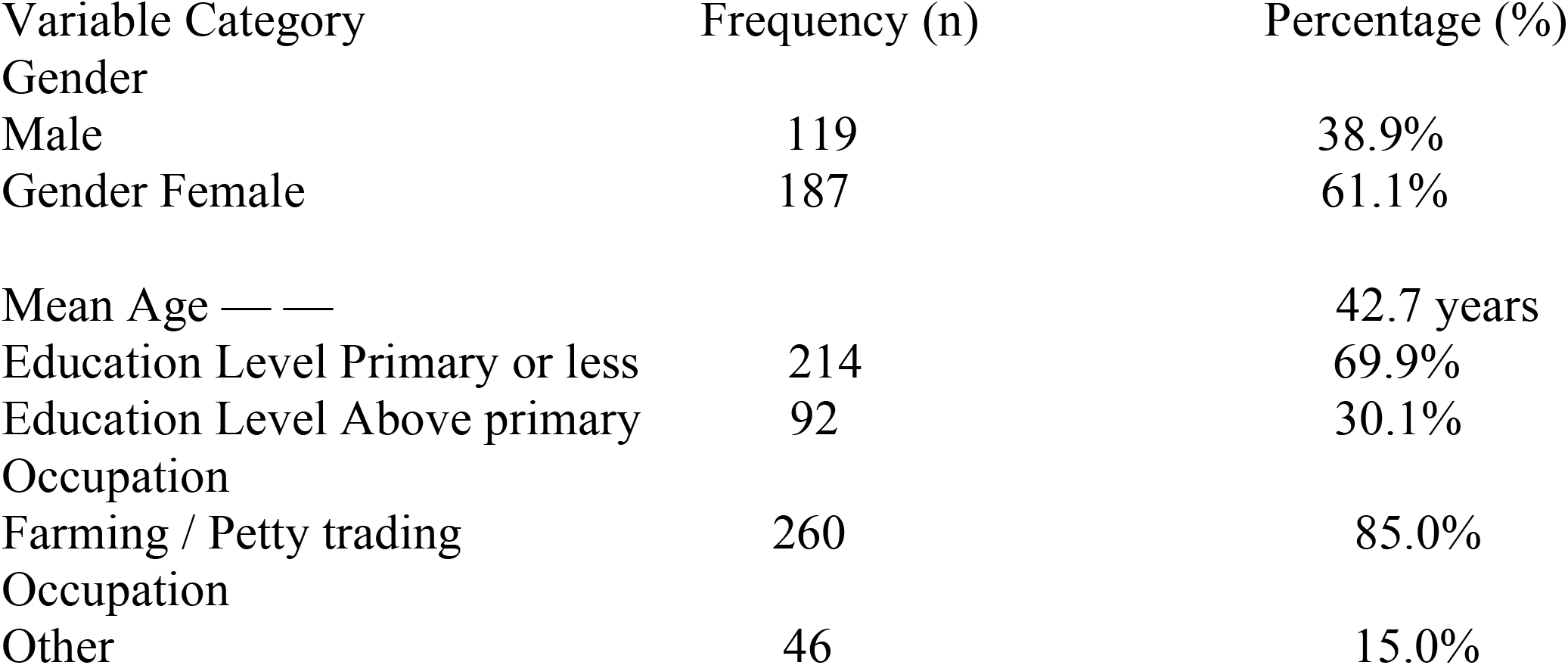
Socio-demographic Characteristics of Respondents (n = 400) Socio-demographic Profile of Respondents (N = 306)

| Variable Category | Frequency (n) | Percentage (%) |
| --- | --- | --- |
| Gender |  |  |
| Male | 119 | 38.9% |
| Gender Female | 187 | 61.1% |
| Mean Age — — |  | 42.7 years |
| Education Level Primary or less | 214 | 69.9% |
| Education Level Above primary | 92 | 30.1% |
| Occupation |  |  |
| Farming / Petty trading | 260 | 85.0% |
| Occupation |  |  |
| Other | 46 | 15.0% |

**Table 2.** Treatment-Seeking Patterns for Buruli Ulcer (N = 306)

| Treatment Behavior | Frequency (n) | Percentage (%) |
| --- | --- | --- |
| First consulted traditional healers | 120 | 30% |
| Sought formal care within first month of symptoms | 50 | 12.5% |
| Reported cost as a barrier | 65 | 16.5% |
| Reported distance to clinic as a barrier | 75 | 18.75% |
| Believed BU had a spiritual cause | 90 | 22.5% |

**Table 3.** Knowledge and Beliefs about Buruli Ulcer.

| Knowledge/Belief Statement | Frequency (n) | Percentage (%) |
| --- | --- | --- |
| Recognized BU symptoms (e.g., swelling, ulcers) | 233 | 58.25% |
| Aware that BU is caused by bacteria | 78 | 19.5% |
| Believed BU is caused by curses, ancestral punishment, etc. | 187 | 46.75% |

**Table 4.** Treatment Pathway and Lesion Severity.

| Treatment Type | Category I | Category II | Category III |
| --- | --- | --- | --- |
| Orthodox (n=144) | 85(59.0%) | 45 (31.3%) | 14 (9.7%) |
| Unorthodox (n=256) | 20 (7.8%) | 105 (41.0%) | 131 (51.2%) |

Chi-square test: $p < 0.01$ , indicating a statistically significant association between treatment type and lesion severity at presentation.

**Table 5.**
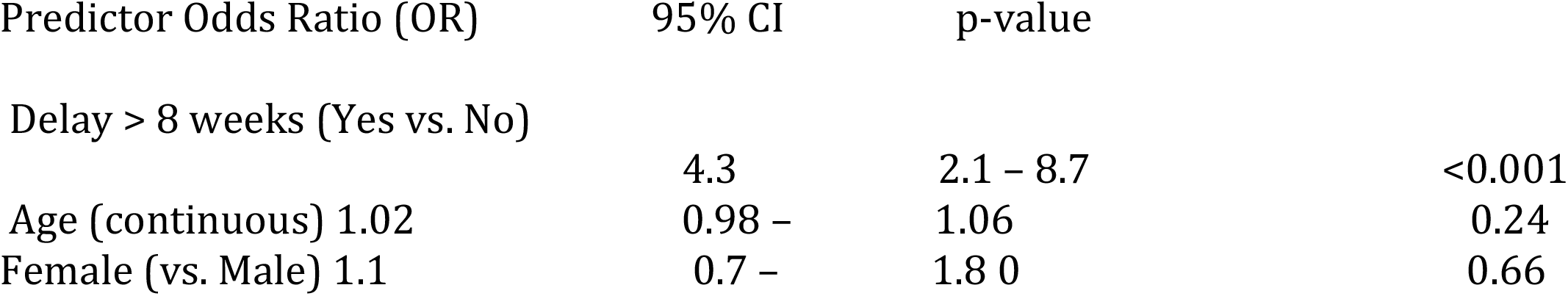
Logistic Regression for Predictors of Late Presentation (Category III Lesions)

**Table 6.**
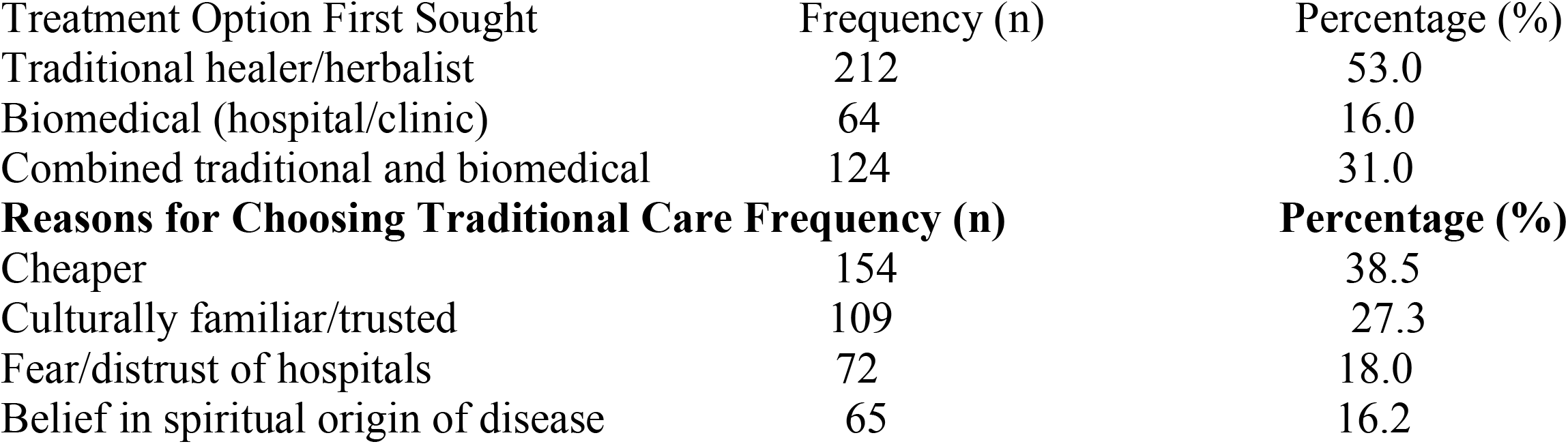
Treatment Preferences and Practices.

| Treatment Option First Sought | Frequency (n) | Percentage (%) |
| --- | --- | --- |
| Traditional healer/herbalist | 212 | 53.0 |
| Biomedical (hospital/clinic) | 64 | 16.0 |
| Combined traditional and biomedical | 124 | 31.0 |
| **Reasons for Choosing Traditional Care** | **Frequency (n)** | **Percentage (%)** |
| Cheaper | 154 | 38.5 |
| Culturally familiar/trusted | 109 | 27.3 |
| Fear/distrust of hospitals | 72 | 18.0 |
| Belief in spiritual origin of disease | 65 | 16.2 |

**Table 7.**
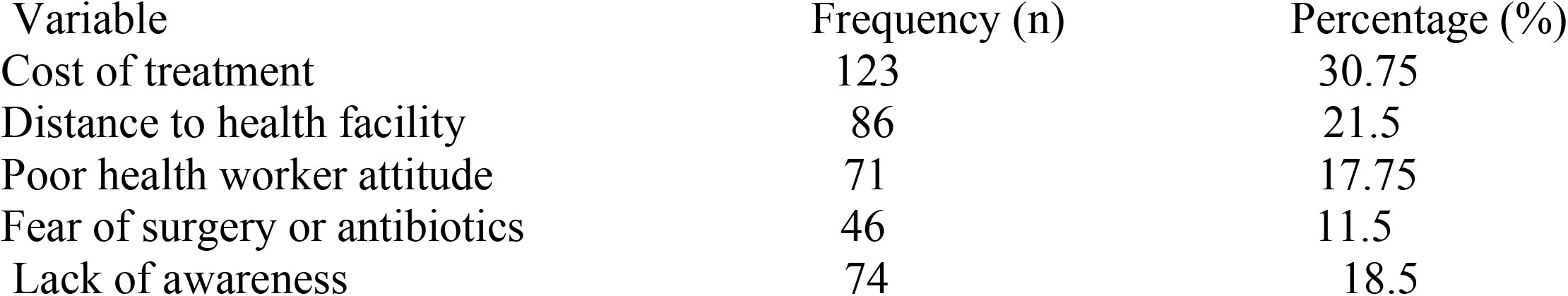
Barriers to Biomedical Treatment Barrier.

**Table 8.** Summary of Key Themes from Qualitative Data Theme Description.

| Theme | Description | Representative Quote |
| --- | --- | --- |
| Cultural interpretations of BU | BU is seen as a spiritual affliction or ancestral curse. | *"We believe this sore is a punishment from the gods or a sign that someone has offended the ancestors."* – FGD, Oguta |
| Efficacy of traditional treatments | Herbal treatments perceived to reduce swelling/pain. | *"Herbs help to dry the sore and make it less painful, but sometimes it comes back."* – Traditional healer, Obowo |
| Barriers to biomedical treatment | Financial cost, distance, fears of hospital care. | *"By the time they reach the hospital, the wound is already big. Transport is a problem."* – Community Elder, Ikeduru |
| Suggestions for integration | Calls for collaboration between health workers and traditional healers. | *"If doctors can work with our native doctors, people will come out early."* – Health worker, Ikeduru |

### 3.2 Qualitative Results

#### Thematic analysis of interviews and focus groups revealed four major themes

Figure 1. : Thematic Map of BU Treatment Dynamics (See diagram below)

##### Theme 1: Cultural Attribution of BU

Many participants believed BU was caused by witchcraft or curses. Traditional healers were seen as the first line of defense due to their spiritual insight. *“People say it comes from enemies or evil spirits… that’s why they go to native doctors first*.*”*

##### Theme 2: Economic Barriers to Orthodox Care

Financial constraints were a major factor in delaying biomedical treatment. The cost of consultation, transportation, and medication deterred early hospital visits. *“Hospitals ask for money we don’t have, but the herbalist is near and cheaper*.*”*

##### Theme 3: Trust in Traditional Healers

There is a deep-rooted trust in traditional practitioners, who are perceived as more empathetic and community-oriented.

##### Theme 4: Willingness to Integrate Treatment

Several participants suggested a collaborative model between orthodox and indigenous healers to improve early treatment uptake. *“If doctors and native healers work together, maybe treatment will be faster*.*”*

**Fig. 3_2.**
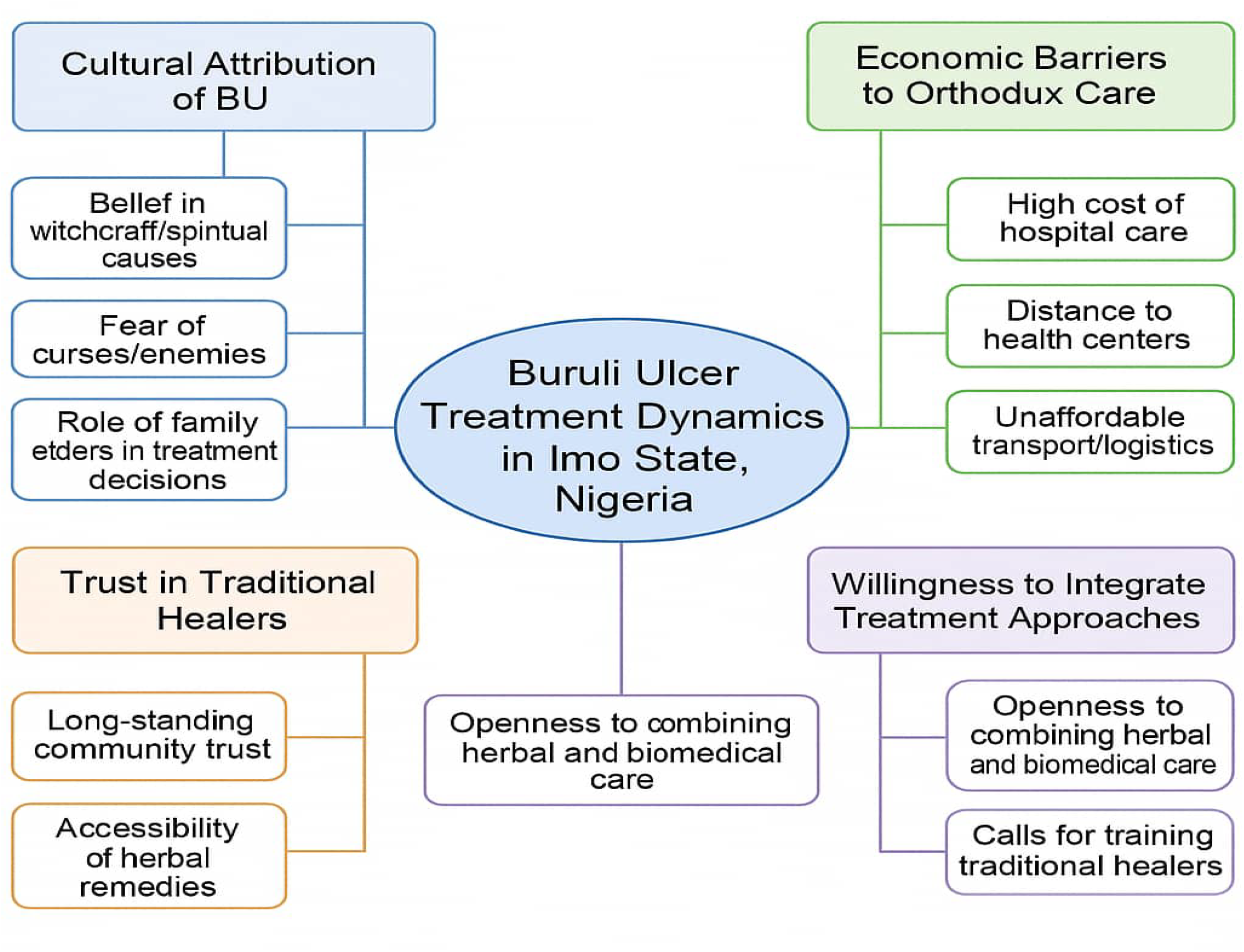
Thematic Map of BU Treatment Dynamics

**Fig. 3_2.**
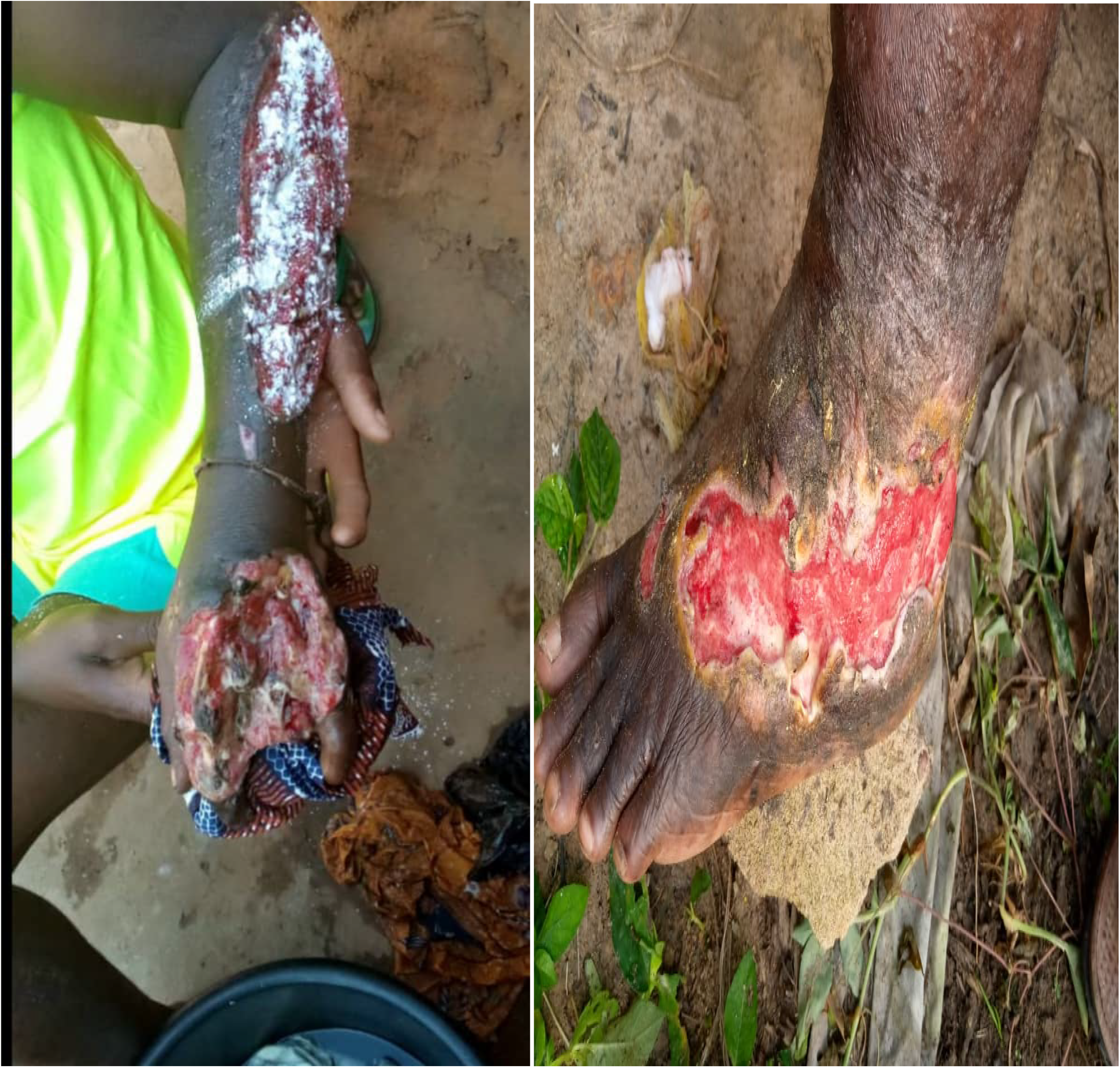
(A) Indigenous Treatment with talisman (B) Biomedical Healing of a Respondent

## 4. DISCUSSION

### Integration of Quantitative and Qualitative Findings

The integration of results highlighted the reinforcing relationship between belief systems and health behaviors. Patients’ cultural understanding of illness and the economic landscape directly influenced their treatment-seeking choices and outcomes. The overlap between statistical trends and thematic narratives supports the need for a hybrid care model. Median delay before orthodox care was 12 weeks (IQR: 8–16). Logistic regression showed patients who delayed orthodox treatment for over 8 weeks had 4.3 times the odds of presenting with Category III lesions (p<0.001).

Factors influencing treatment choice included cultural attribution of disease to witchcraft (40%), economic barriers (35%), and accessibility issues (25%). Qualitative findings highlighted deep community trust in traditional healers, cost-related avoidance of hospitals, and interest in collaborative care models. Thematic mapping revealed four key themes: (1) Cultural Attribution of BU, (2) Economic Barriers to Orthodox Care, (3) Trust in Traditional Healers, and (4) Willingness to Integrate Care Systems. The findings of this study offer critical insights into the complex socio-cultural and economic landscape that shapes Buruli Ulcer (BU) treatment decisions in Imo State, Nigeria.

The high rate (64%) of initial reliance on traditional healing practices is consistent with previous research in West Africa, where cultural interpretations of illness and accessibility challenges strongly influence health-seeking behaviors (Garchitorena et al., 2019; Adebayo et al., 2022). One of the dominant themes emerging from this study is the attribution of BU to supernatural causes such as witchcraft, spiritual attacks, or curses. This aligns with literature indicating that in many African communities, biomedical explanations of disease are often secondary to spiritual and metaphysical interpretations (Yeboah-Manu et al., 2021).

Consequently, traditional healers, who are embedded within community structures and perceived to possess esoteric knowledge, serve as the first point of care. Economic barriers also significantly affect healthcare choices. Our findings show that costs associated with hospital visits, medication, and transportation deters early access to orthodox care. This corroborates evidence from the WHO and other studies identifying out-of-pocket expenses as a major barrier to treatment of neglected tropical diseases (WHO, 2023; Nigerian Ministry of Health, 2023). In such contexts, traditional remedies are often preferred due to their affordability and flexible payment systems.

The logistic regression results highlight the clinical implications of delayed care. Patients who waited more than 8 weeks before seeking orthodox treatment were over four times more likely to present with severe (Category III) lesions. This underscores the urgent need for early diagnosis and prompt treatment, both of which are cornerstones of WHO’s BU control strategy (WHO, 2023). Qualitative findings further revealed a promising avenue for intervention: the community’s openness to integrating traditional and biomedical approaches. Participants expressed a willingness to accept a hybrid model where traditional healers could refer complicated cases to biomedical facilities and receive training on recognizing BU symptoms. This mirrors integration efforts in other low-resource settings, where collaboration between health systems and indigenous practitioners has improved health outcomes (Garchitorena et al., 2019).

Overall, these results highlight that while traditional healing systems may delay effective treatment, they also offer a potential bridge to enhance early intervention if properly integrated into public health strategies. Culturally tailored health education, improved healthcare infrastructure, and policy support for traditional practitioner engagement are vital for reducing BU morbidity and mortality in Nigeria. Empowering traditional healers with referral knowledge and training can facilitate early diagnosis and treatment.

## CONCLUSION

Buruli Ulcer treatment dynamics in Imo State are shaped by a complex interplay of cultural, economic, and systemic factors. To mitigate disease burden, a hybrid care approach that respects indigenous beliefs while promoting evidence-based protocols is essential. Policy recommendations include rural health education campaigns, healthcare subsidies, and formalized collaboration with traditional practitioners.

### Recommendations

- Develop culturally tailored health education programs
- Offer financial subsidies for BU treatment and transport
- Train traditional healers on BU symptom recognition and referral

## Data Availability Statement

The data presented in this study are available on request from the corresponding author.

## Author Contributions

Conceptualization: Onwuka Chigozie Divine; Methodology: Oparaocha Evangeline Tochi; Data Collection: Both authors; Formal Analysis: Onwuka Chigozie Divine; Writing – Original Draft: authors; Writing – Review & Editing: Both authors.

## ACKNOWLEDGEMENTS

We thank the Imo State Ministry of Health, participating health facilities, traditional healers, and the study participants for their support.

